# Unveiling Breast Cancer Risk Profiles: A Comprehensive Survival Clustering Analysis Empowered by an Online Web Application for Personalized Medicine

**DOI:** 10.1101/2023.05.18.23290062

**Authors:** Yuan Gu, Mingyue Wang, Yishu Gong, Song Jiang, Chen Li, Dan Zhang

## Abstract

Online tools, such as web-based applications, aid medical doctors in recommending treatments and conducting thorough patient profile investigations. Prior studies have created web-based survival analysis tools for cancer survival. However, these often offer basic features and simplistic models, providing shallow data insights. Our research involves an in-depth risk profile analysis using survival clustering on real-world data. We’ve developed a user-friendly Shiny application to simplify the use of our findings. By utilizing survival clustering, we uncover distinct subgroups and unique risk profiles among breast cancer patients. Our online app empowers researchers and clinicians to explore and gain insights into breast cancer risk profiles, enhancing personalized medicine and clinical decision-making.

## 1. Introduction

Understanding the risk profile and survival outcomes of breast cancer remains a complex and intricate area of research. Despite significant advancements in the field, there are still numerous factors that contribute to the variability in breast cancer progression and patient outcomes. The interplay between genetic predisposition, lifestyle factors, tumor characteristics, and treatment response adds to the challenge of unraveling the intricacies of breast cancer risk profiles. Additionally, the heterogeneity within breast cancer itself further complicates the identification of clear-cut survival patterns. Consequently, there is a pressing need to delve deeper into the complexities of breast cancer risk profiles and survival to uncover new insights and potential avenues for improved diagnosis, treatment, and personalized care for patients.

In this study, we undertake a comprehensive analysis of risk profiles using survival clustering techniques applied to the Molecular Taxonomy of Breast Cancer International Consortium (METABRIC) dataset [1]. An unsupervised learning approach was used to cluster patients based on their survival difference and other relevant clinical variables, by combining k-means clustering and Kaplan-Meier (K-M) curves, leveraging the significant risk factors identified through a Cox regression model. Through a deeper exploration of survival clustering, we can discover heterogeneous subpopulations with varying survival characteristics including age, tumor stage and molecular subtypes etc., providing insights into the underlying heterogeneity of breast cancer and reveal potential biomarkers for risk stratification. Additionally, we have developed an accessible online application using Shiny to facilitate easier utilization of our findings.

Currently, there is a scarcity of web-based survival analysis tools available for medical researchers conducting survival studies and all previous web applications have primarily focused on Cox regression [2] or genetic analysis and other data structures [3–6]. Also, a lot of previous relevant studies used semi-supervised or self-supervised learning for medical multi classification problems but lack of survival information [7–8]. In contrast, our study fills an existing void by introducing a unique combination of unsupervised learning techniques and risk factor analysis on the clinician side, demonstrating the potential of survival clustering as a valuable tool in uncovering hidden structures based on distinct risk profiles. Overall, our online tool provides a user-friendly interface for researchers and clinicians to explore the results and derive valuable insights on breast cancer. The link for this app is: https://baran-shad.shinyapps.io/breastcancer

## 2. Literature review

Breast cancer is a complex disease with diverse clinical outcomes, making accurate prediction of survival crucial for personalized treatment and care. Over the years, various survival models have been developed to aid in understanding and predicting cancer survival rates, from various perspectives, such as increasing the prediction accuracy from the model aspect by using metrics like AUC and AUC-PR [9]. The most widely used classical approach is the Cox regression model, which assumes proportional hazard rate. Numerous studies have applied this model to identifying significant prognostic factors such as age, tumor stage, hormone receptor status, and lymph node involvement [10–11]. However, the Cox model relies on the proportional hazard assumption, which may not always hold true, leading to potential bias in the estimation of survival probabilities.

To address the limitations, researchers have explored alternative techniques, including machine learning algorithms, such as random forests, support vector machines, and neural networks, which have demonstrated promising results [12–15]. These models can handle complex interactions between variables and capture non-linear relationships, thus providing improved accuracy in survival prediction. However, their black-box nature often limits interpretability and understanding of the underlying biological mechanisms.

Another noteworthy approach is the use of gene expression data. Gene expression-based models, such as using multi-omics neural networks to make the survival prediction, have shown the ability to provide deeper insight into which types of data are most relevant to improve prognosis [16–17]. These models provide valuable insights into the underlying biology of breast cancer and offer potential for personalized treatment strategies. However, their reliance on gene expression data may limit their application in clinical settings where gene expression profiling is not routinely performed. Other challenges and limitations exist in the data and visualization, our previous sickle cell disease studies also combined machine learning and survival model but without visualization interactive online tools [18, 19]. Issues like missing data, limited sample sizes, and the lack of accessible visualization tools for physicians and medical professionals with limited modeling expertise hinder the widespread utilization of the models.

In our study, we address all above challenges by developing a user-friendly interface to uncover hidden structures within breast cancer data and identify unique risk profiles. This intuitive tool enables researchers and clinicians to easily explore and interpret the results of our analysis and to gain valuable insights into the complex landscape of breast cancer risk profiles, bridging the gap between sophisticated modeling techniques and practical clinical applications.

## 3. Methods

### 3.1 Data

The METABRIC dataset is a valuable and publicly accessible resource for researchers. It encompasses a total of 2,506 subjects and 34 variables, providing a comprehensive foundation for studying breast cancer. To ensure the reliability of our analysis, we diligently handled any missing data and performed thorough feature checks. A total of 528 subjects who lacked survival status information were excluded from the analysis. After removing the 528 subjects, other certain variables with limited variability, such as Sex that comprised all females, were also omitted from the dataset. Following the above meticulous data preprocessing procedures, our final analysis dataset consists of 1,269 subjects and 23 variables without missing values. The 23 variables are : Age at diagnosis, Subtype cohort (termed integrative clusters )[20], neoplasm histologic grade, number of lymph nodes examined to be positive, mutation count, Nottingham prognostic index (NPI), overall survival time in months, relapse free status in months, tumor size, tumor stage, cancer type, ER status, HER2 status, hormone therapy status, survival status, prior radiology status, menopausal status, integrative subgroup, chemotherapy, cellularity, Prediction Analysis of Microarray 50 (PAM50), relapse status. All analysis is built by R (v 4.3.0). The descriptive statistics, including means and 95% confidence intervals for continuous variables and count and proportions for categorical variables, for the 23 variables in each survival status group (living or decreased) are provided in the following Table 1. A t-test was used to compare the Age at diagnosis between the survival status groups (living versus decreased), and a Chi-Square test was used to examine the categorical variables between the survival status groups (living versus deceased). The variables with p-value <0.05 suggest a statistically significant difference between the groups.

**Table 1:** Basic descriptive statistics. * p value denotes significant level when p<0.05 ** continuous variables are summarized by mean (min-max)

|  | Living(N=548) | Deceased (N= 721) | p-value |
| --- | --- | --- | --- |
| Age.at Diagnosis** | 56 (26.72- 85.21) | 66 (21.93- 96.29) | <0.0001 * |
| Tumor Size | 23 (1 - 100) | 29 (1 - 180) | <0.0001 * |
| Neoplasm Histologic Grade |  |  | <0.0001 * |
| 1 | 55(10.04%) | 46 (6.38%) |  |
| 2 | 228 (41.61%) | 269(37.31) |  |
| 3 | 265 (48.36%) | 406(56.31%) |  |
| Lymph nodes examined positive | 1.15(0-25) | 2.45(0-41) | <0.0001* |
| Mutation Count | 4.9 (1-26) | 5.9(1-46) | <0.0001* |
| Nottingham prognostic index | 3.9(2-6.19) | 4.3(2-6.36) | <0.0001* |
| Tumor Stage |  |  | <0.0001* |
| 1 | 227(41.42%) | 193(26.77%) |  |
| 2 | 293 (53.47%) | 443(61.44%) |  |
| 3 | 27(4.93%) | 78(10.82%) |  |
| 4 | 1(0.18%) | 7(0.97%) |  |
| ER Status |  |  | 0.54 |
| Negative | 121(22.08%) | 170(23.58%) |  |
| Positive | 427(77.92%) | 551(76.54%) |  |
| HER2 Status |  |  | 0.037* |
| Negative | 494(90.15%) | 622(86.27%) |  |
| Positive | 54(9.85%) | 99(13.73%) |  |
| Hormone therapy |  |  | 0.39 |
| No | 227(41.42%) | 281(38.97%) |  |
| Yes | 321(58.58%) | 440(61.03%) |  |
| PR Status |  |  | 0.023* |
| Negative | 242(44.16%) | 365(50.62%) |  |
| Positive | 306(55.84%) | 356(49.38%) |  |
| Relapse Status |  |  | <0.0001* |
| Not Recurred | 479(87.41%) | 255(35.37%) |  |
| Recurred | 69(12.59%) | 466(64.63%) |  |
| Menopausal |  |  | <0.0001* |
| Pre | 171(31.20%) | 122(16.92%) |  |
| Post | 377(68.80%) | 599(83.08%) |  |
| Integrative number | 6.30(1-11) | 6.32(1-11) | 0.89 |
| Chemotherapy |  |  | 0.12 |
| No | 416(75.91%) | 574(79.61%) |  |
| Yes | 132(24.09%) | 147(20.39%) |  |
| Cellularity |  |  | 0.54 |
| Low | 65 (12%) | 76 (11%) |  |
| Moderate | 218(40%) | 275(38%) |  |
| High | 265(48%) | 370(51%) |  |
| Pam50Claudin | 3.80(1-7) | 3.86(1-7) | 0.35 |

### 3.2 Model

The analysis of the breast cancer data encompassed three distinct phases. In the first phase, Cox regression with stepwise AIC selection was used to identify statistically significant risk factors. This approach allowed us to determine the variables that most significantly influenced breast cancer outcomes among the 22 variables. Following this, K-means clustering was conducted based on the selected risk factors from the Cox model in the first phase. By grouping similar individuals together, this clustering analysis provided insights into distinct subgroups within the dataset. In the final phase, a Kaplan-Meier model was constructed using the predicted clusters, enabling a deeper exploration of the risk profiles associated with each cluster. This comprehensive approach allowed for a thorough examination of the relationship between risk factors, clustering patterns, and breast cancer outcomes, ultimately enhancing our understanding of the disease.

The first phase is the Cox regression with stepwise AIC selection. Cox regression, assumes proportional hazards, is specifically designed for survival analysis, allows us to assess the impact of various variables on the time until death occurs. By using stepwise AIC selection, the model identifies the subset of variables that provide the best fit for the data, while controlling for the risk of overfitting. This approach considers the trade-off between model complexity and goodness of fit, selecting a parsimonious model that optimizes the AIC criterion.

Once significant risk factors were identified, the next phase is the k-means clustering to group individuals based on those factors. To determine the optimal number of clusters, various methods such as the elbow method, Silhouette coefficient, and gap statistics were considered as the standard methods in previous research. However, because K-means clustering is unsupervised learning, without label information, the optimal number of clusters determination is often challenging and subjective. Choosing an inappropriate value for k can lead to very poor clustering results, for example, if a dataset contains some outliers which can significantly affect the position and size of the clusters, the above widely used methods may provide incorrect clustering assignments. In that case, preprocessing steps or outlier detection techniques may be needed to mitigate this issue, however, in survival data, one event outlier might be very informative compared to other normal data, indicating a person who has higher risk to get the event. Given that our ultimate objective was to examine the survival risk profile, in the final phase, we incorporated the Kaplan-Meier (KM) model, the log-rank test, to identify differences between survival curves among clusters. By leveraging the insights provided by the log-rank test, we are able to select a different number of clusters and visualize the results. The visualized clusters were also presented in the web-app, allowing for interactive exploration.

For each number of clusters, we conducted an analysis of the basic characteristics associated with the predicted clusters. By assessing these characteristics, we gained insights into the distribution and patterns within each cluster. For continuous risk variables, the mean values provided an indication of the average risk level within that cluster. Meanwhile, for categorical risk factors, the frequency analysis allowed us to identify the prevalence of specific risk factors within each cluster. This comprehensive examination of basic characteristics facilitated a deeper understanding of the distinct profiles.

## 4. Results

The Cox regression analysis with AIC selection identified several significant risk factors associated with breast cancer outcomes; the result is presented in Table 2.

**Table 2:** Cox regression results.

| Predictors | Survival Model |  |
| --- | --- | --- |
|  | Estimates | p-value |
| Age at diagnosis | 0.04 | <0.01 |
| Cohort | 0.07 | 0.12 |
| Mutation Count | 0.02 | 0.08 |
| Nottingham prognostic index | 0.12 | 0.01 |
| Relapse Free Status Months | -0.02 | <0.001 |
| Integrative number | 0.02 | 0.11 |
| Tumor Stage | 0.12 | 0.11 |
| ER Status | -0.56 | <0.001 |
| HER2 Status | 0.21 | 0.07 |
| Hormone Therapy | 0.21 | 0.04 |
| Chemotherapy | 0.3 | 0.03 |

A total of 6 variables are statistically significant risk factors. Age at diagnosis (p <0.01) showed a significant positive relationship with death, suggesting that older age is associated with increased risk. A higher Nottingham Prognostic (p = 0.01) Index, relapse-free periods (p-value < 0.001), ER-negative status (p < 0.001) and experiencing Hormone and Chemo therapies are also associated with increased hazard rates.

The model has a concordance of 0.897, demonstrating a very strong discriminatory power to distinguish between individuals with different survival outcomes with a high degree of accuracy. In the next step, all the above significant risk factors are standardized by simply converting into a z-score and then used as the input variables into the k-means clustering, to ensure all input variables are on the same scale, preventing variables with larger magnitudes from dominating the clustering process. In this phase, K-means will be used to partition the dataset into K clusters. The algorithm assigns each data point to the cluster with the nearest mean value.

Although traditional k-means is unsupervised learning without predefined labels or target variables, in our case, we improved it by incorporating a pseudo-supervised way, such as the K-means survival difference. We used the survival difference between different clusters to visually guide the selection of the appropriate number of clusters. This approach adds an extra layer of information beyond the traditional K-means, enabling the model to assess the quality of clustering based on survival differences.

Suppose we assign K number of clusters to all observations and the k clusters are denoted □_□_ □□□□□□ = *1*… □, the log rank statistic is approximately distributed as a chi-square test statistic, and the following is the test formula (1):

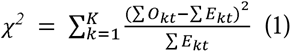

where ∑*O_kt_* represents the sum of the observed number of events in the □^□□^ cluster over time, and ∑*E_kt_* represents the sum of the expected number of events in the □^□□^ cluster over time. and the K-means cluster algorithm is by solving the following optimization problem (2):

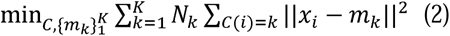

where □(□) = □ stands for assigning the ith observation to the □^□□^ cluster C, □_□_ is the mean value of □^□□^ cluster. □_□_ are the observations in the □^□□^ cluster. The objective of the algorithm is to minimize the total dissimilarity by assigning N observations to K clusters in a manner that minimizes the average dissimilarity of the observations from their respective cluster means. The dissimilarity is calculated by the 2-norm stand for ‖·‖*^2^*. Our analysis and the web application are based on incorporating both (1) and (2) for different K assignments. The following Picture 1 and 2 presents the clustering points and the survival curve difference by selecting k = 4 and 5, more other k choices are available in the web application.

**Picture 1.**
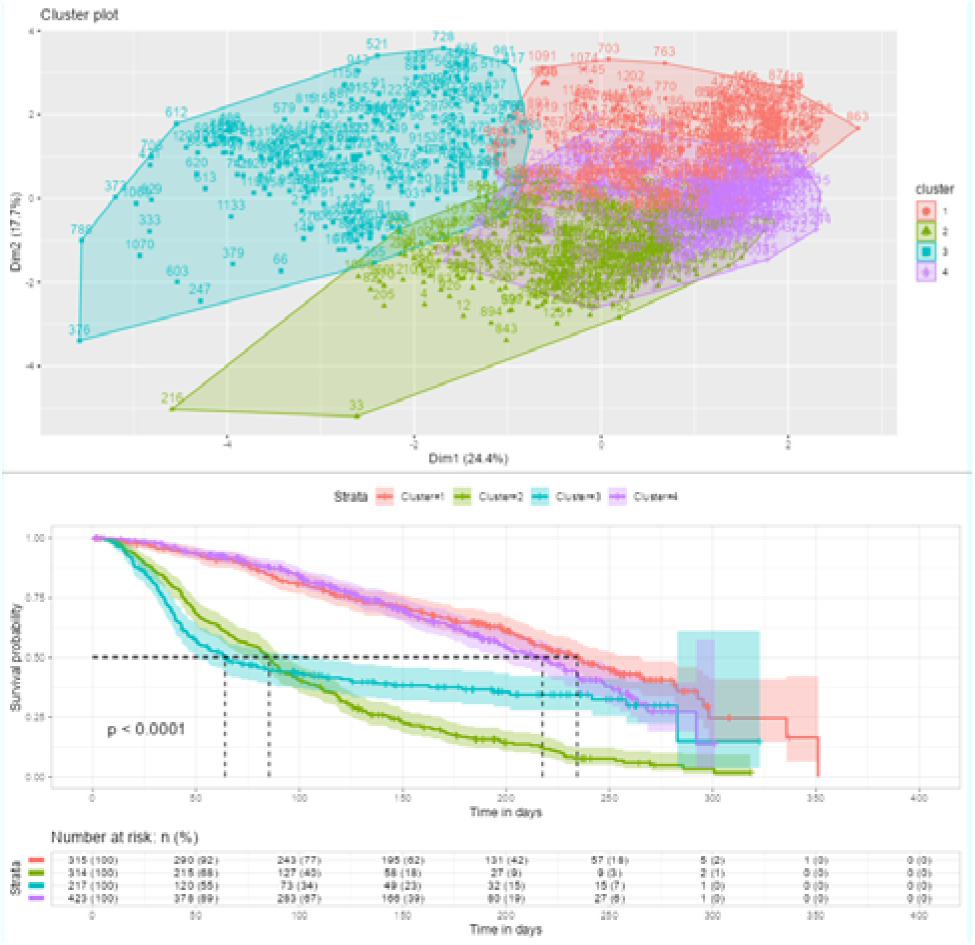
4 clusters and the K-M curves

**Picture 2.**
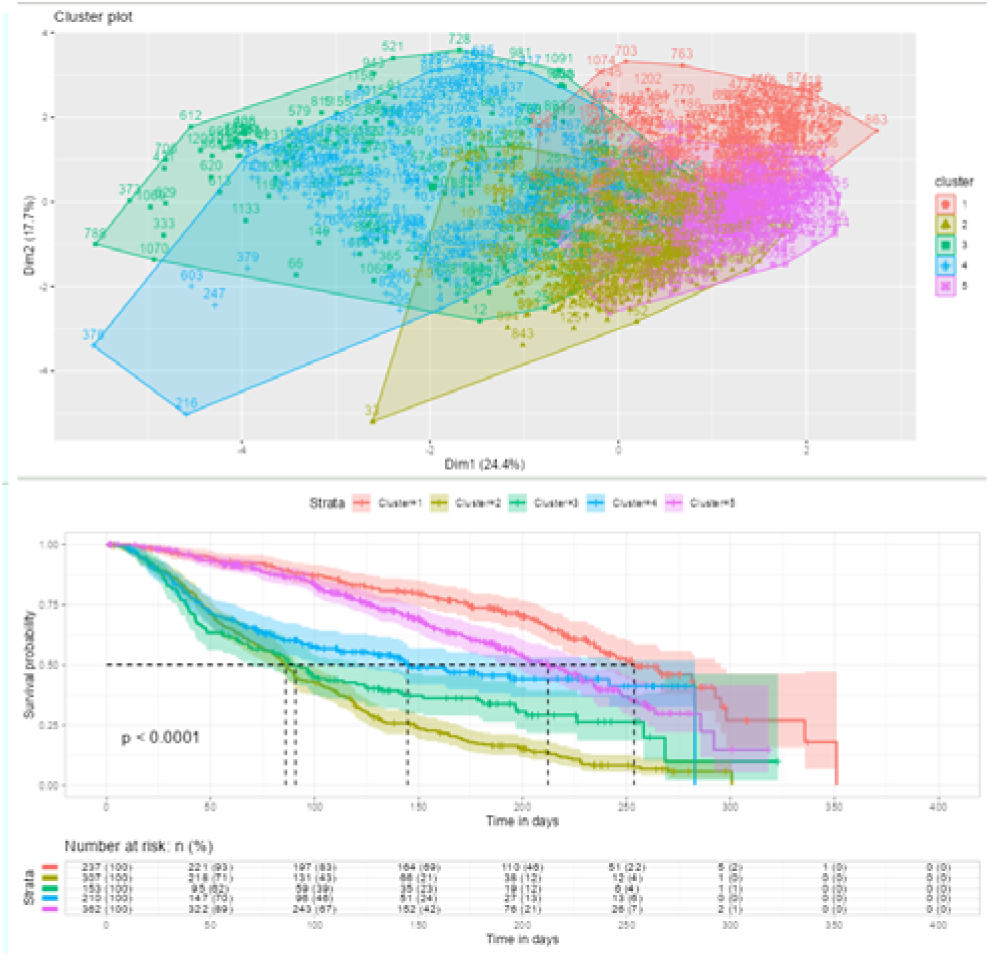
5 clusters and the K-M curves

In Picture 1, four distinct clusters are discernible, each highlighted with a unique color (upper). The accompanying graph below represents the survival plot (below). The colors corresponding to clusters 1 to 4 are red, green, blue and purple. The sample sizes of these clusters vary, with 315, 314, 217 and 423 from cluster 1 to 4. Significant survival differences can be observed when comparing clusters (1 and 4) to clusters (2 and 3). Additionally, within these groupings, minor survival differences are also evident between clusters 1 and 4, as well as between clusters 2 and 3. However, in Picture 2, when we select k = 5, per reconfiguration, we now observe the colors for clusters 1 to 5 are red, yellow, green blue and purple, also clusters 1 and 5, very close to the former clusters 2 and 3 when k was set to 4. Notably, there is a significant survival difference between these two clusters (1 and 5) compared to the remaining three clusters (clusters 2, 3, and 4). In terms of sample size, clusters 1 through 5 consist of 237, 307, 153, 210, and 362 samples, respectively. The p-values obtained from the log-rank test are both (k=4 and k=5) less than 0.0001, as indicated in the accompanying survival plots. The median survival time, represented by the horizontal dashed line in the plots, is also added in the survival plots.

Our final phase involves a comprehensive examination of the risk profiles across the different clusters. After assigning clusters, we can delve deeper into those significant risk factors. Table 3 shows the details for continuous risk factors for 4 clusters when K=4. Picture 3 is the box plot for continuous risk factors by 4 clusters and this illustration provides more specific details in the web application. Furthermore, we also depicted the frequency distribution of each subgroup within categorical variables (e.g., recurrence vs. no recurrence for the ’Relapse Status’) stratified by clusters in the following illustration (Picture 4).

**Picture 3.**
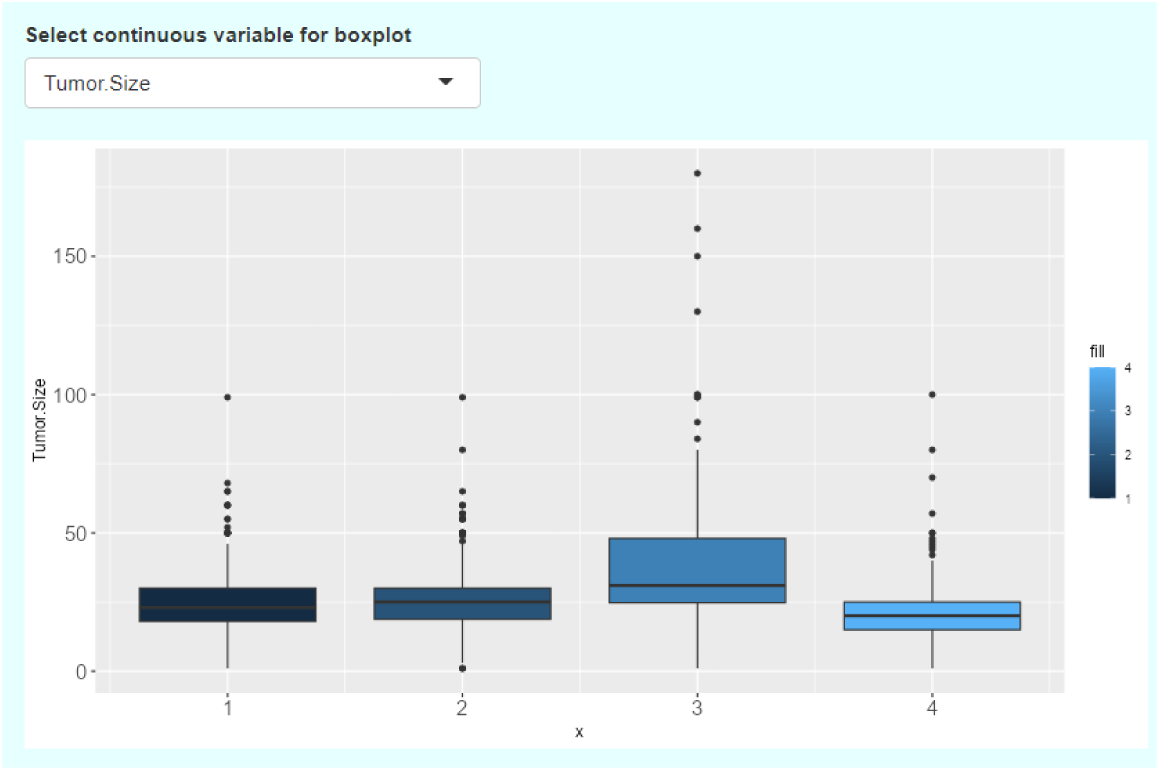
Box plot for Tumor size group by 4 clusters

**Picture 4.**
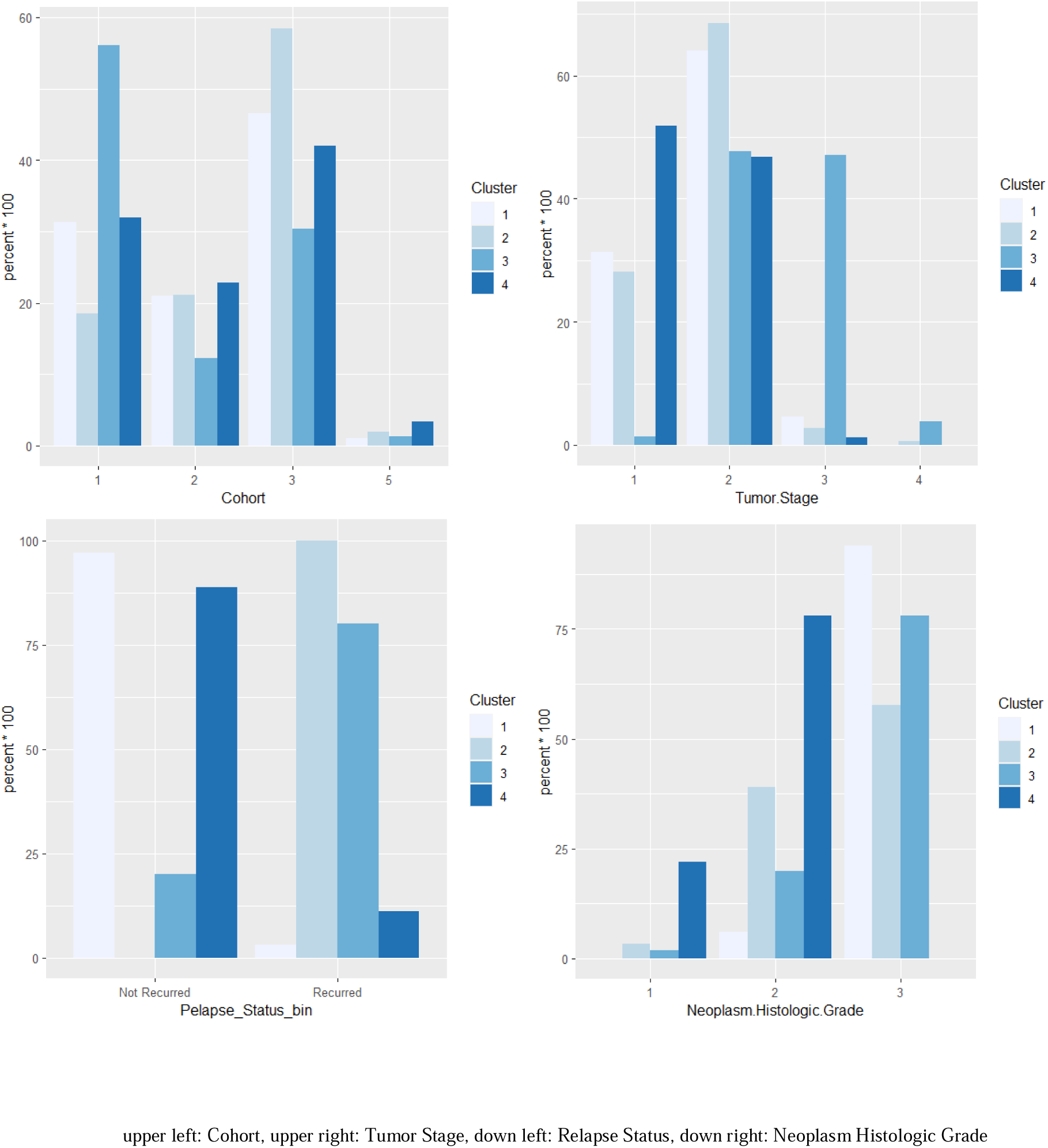
Categorical subgroup frequency group by 4 clusters

**Table 3:** Summary Statistics for continuous risk factor by 4 clusters.

| Cluster (n) | Number of Events | Median Survival time in months | Age at diagnosis (range) | Nottingham prognostic index (range) | mutation counts (range) |
| --- | --- | --- | --- | --- | --- |
| 1 (n=367) | 111 | 260.0 | 59.73<br>(26.72-90.08) | 4.51<br>(3.05-6.08) | 5.55<br>(1-40) |
| 2 (n=356) | 344 | 75.0 | 59.91<br>(26.36-90.23) | 4.14<br>(2.03-6.07) | 5.92<br>(1-30) |
| 3 (n=155) | 131 | 46.1 | 61.15<br>(21.93-96.29) | 5.72<br>(4.03-6.36) | 4.83<br>(1-22) |
| 4 (n=391) | 135 | 251.2 | 61.08<br>(33.76-90.43) | 3.12<br>(2 - 5.06) | 5.27<br>(1-46) |

In the provided table and pictures, we observe that cluster 3 exhibits the shortest median survival time (Table 3) and the largest tumor size (Picture 3). Additionally, in the frequency plot of tumor stage, cluster 3 comprises the highest number of patients with stage 3 and above. Furthermore, the Neoplasm Histologic Grade in cluster 3 is also the highest compared to the other cluster groups. Cluster 2 also demonstrates a relatively lower survival rate compared to the other clusters. From Picture 4, we can discern a pattern where cluster 2 has higher Neoplasm Histologic Grade and tumor stages compared to clusters 1 and 4.

Upon setting K = 5, it can be observed that cluster 1 and cluster 5 exhibit lower survival rates in the following Table 4 and Picture 5. Moreover, these two clusters are characterized by relatively higher Neoplasm Histologic Grade compared to the other clusters. In terms of relapse status, cluster 1 comprises the majority of patients who have not experienced recurrence, whereas cluster 5 has the highest number of patients with recurrence. These findings indicate the need for further investigation from a clinical perspective to gain deeper insights.

**Picture 5.**
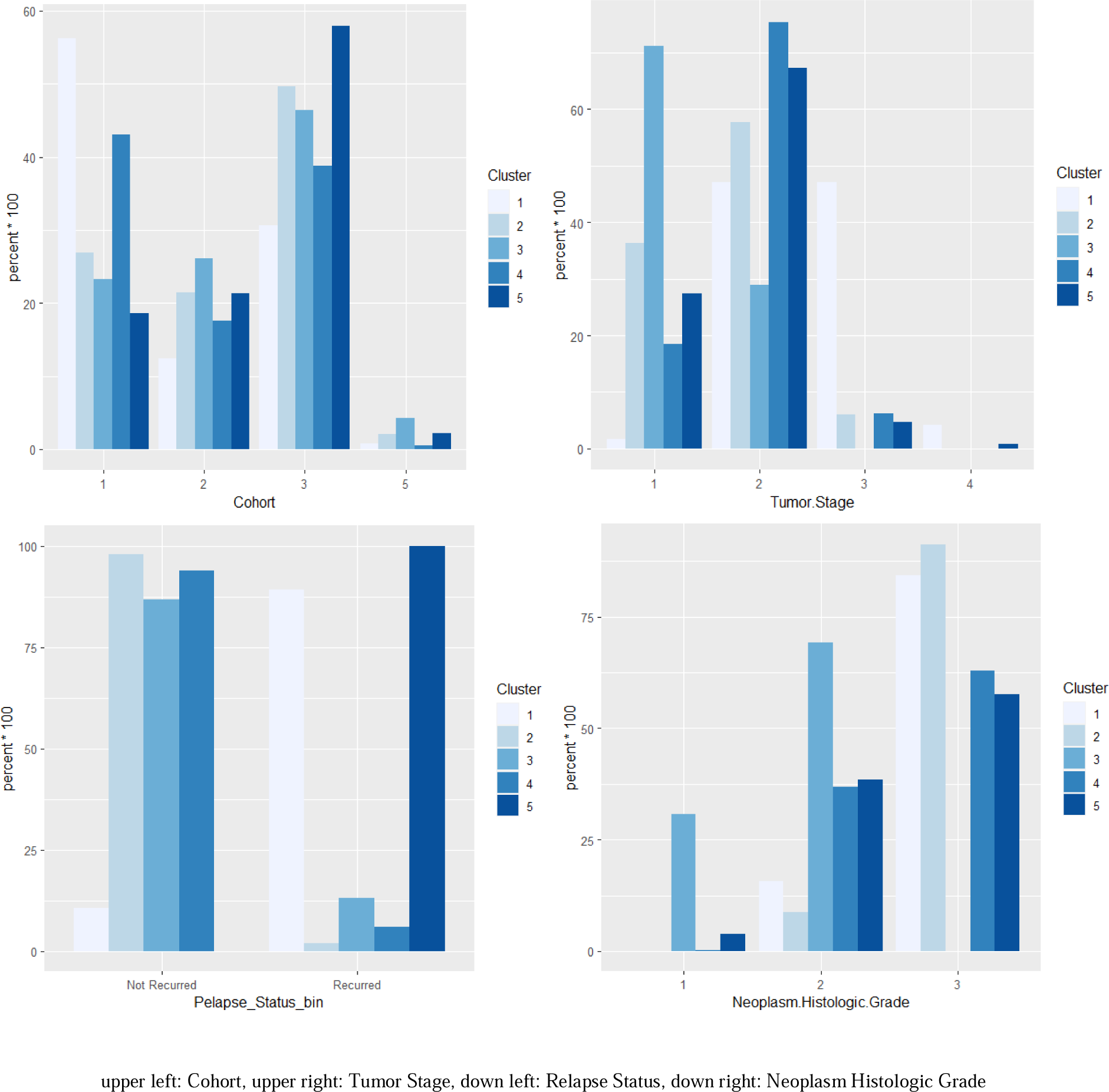
Categorical subgroup frequency group by 5 clusters

**Table 4:** Summary Statistics for continuous risk factor by 5 clusters.

| Cluster (n) | Number of Events | Median Survival time in months | Age at diagnosis (range) | Nottingham prognostic index (range) | mutation counts (range) |
| --- | --- | --- | --- | --- | --- |
| 1 (n=121) | 107 | 43.2 | 59.73 (26.72-90.08) | 4.514 (3.05-6.08) | 5.55 (1-40) |
| 2 (n=149) | 33 | 282.8 | 59.91 (26.36-90.23) | 4.144 (2.03 – 6.07) | 5.92 (1-30) |
| 3 (n=280) | 91 | 298.0 | 61.15 (21.93 – 96.29) | 5.719 (4.03- 6.36) | 4.83 (1-22) |
| 4 (n=353) | 133 | 227.9 | 61.08 (33.76 – 90.43) | 3.122 (2 -5.06) | 5.274 (1-46) |
| 5 (n=366) | 357 | 71.4 | 60.06 (26.36 – 90.23) | 4.167 (2.03 – 6.08) | 5.956 (1-30) |

## 5. Conclusion and Future Work

In conclusion, our comprehensive analysis consisting of three phases, which involved incorporating survival information into the unsupervised machine learning K-means clustering model and developing a web app, has showcased significant advantages, creativity, and contributions in the analysis of breast cancer data. Through a comprehensive three-phase approach, we have provided valuable insights into the risk factors, clustering patterns, and outcomes associated with breast cancer. Our advantage lies in using Cox regression with stepwise AIC selection as the first phase of analysis. The cox model identifies statistically significant risk factors for breast cancer with a very promising concordance value of 0.937. The second phase involves k-means clustering, which groups individuals based on the selected risk factors from the Cox model. By identifying similar individuals within the dataset, this clustering analysis reveals distinct subgroups and provides a deeper understanding of the data. We consider the optimal number of clusters by log rank test according to the KM model to explore the risk profiles associated with each cluster in the last phase. This in-depth examination enhances our knowledge of the distinct profiles and risk factors associated with each predicted cluster, ultimately contributing to our understanding of breast cancer. In summary, our web app empowers healthcare professionals and researchers to make informed decisions and advance their knowledge in the fight against breast cancer.

However, additional research is necessary to achieve a broader application. The current app is designed specifically for this dataset and lacks a comprehensive investigation of generalizability. To enhance usability for medical professionals, it would be beneficial to develop a more convenient pipeline that allows users to import and download their own datasets and select their own potential risk factors. Another limitation is the relatively small number of input variables in this dataset. Therefore, the development of additional models or tools capable of handling larger datasets should be considered for future endeavors.

## Data Availability

All data produced are available online at

https://www.cbioportal.org/study/clinicalData?id=brca_metabric

